# Home Care Follow up determine the point of inversion of IL-6 levels in relation to C-Reactive Protein as the cytokine storm marker in COVID-19

**DOI:** 10.1101/2022.03.21.22270828

**Authors:** Sérgio Paulo de Mello Mendes Filho, Fernanda Simão Martins, Paulo Jose Giroldi, Raul Honorato e Melo, Edcleia Lopes de Oliveira, Anibal Borin dos Santos, Dayse Cristina Oliveira Medeiros, Sergio de Almeida Basano, Jéssica Amaral Lopes, Yury Oliveira Chaves, Rebeca Sousa Pinheiro, Luís Marcelo Aranha Camargo, Juliana Pavan Zuliani, Paulo Afonso Nogueira

## Abstract

The IL-6 has been used for the characterization of the cytokine storm induced by SARS-CoV-2, but so far, no one has found out when and in whom the cytokine storm develops. Our study demonstrates how early and longitudinal clinical-based monitoring and dosing of five markers (C-reactive protein, IL-6, fibrinogen, ferritin and D-dimer) helped to identify who’d developed the cytokine storm. The peak of IL-6 in pg/mL proportionally higher than the peak of CRP in mg/L was sufficient to define the timing of the evolution of cytokine storm syndrome. The administration of antibiotic therapy, anticoagulant therapy and pulse therapy resolved the infection and prevented the progressive deterioration of the lung function of the patients with potential for development of severe COVID-19.

## Introduction

The evolution to severe acute respiratory syndrome (SARS) in patients with severe COVID-19 is triggered by a remarkable and uncontrolled lung inflammation characterized by a systemic increase in cytokines and chemokines, known as the macrophage activation syndrome (MAS) or more usually called a “cytokine storm” [1–5]. MAS can directly induce tissue damage, coagulopathy, dyspnea, hypoxemia, hypotension and hemostatic imbalance, which induces a necessity of mechanical ventilation and increases the risk of death [2,3,6–10].

A virus-induced cytokine storm includes fever, coagulopathy and immune cell-mediated responses, sharp elevations in acute phase responses [characterized by increases in interleukin 6 (IL-6), ferritin, and C-reactive protein (CRP)] [9,11–17].

Several studies have addressed that in some cases COVID-19 patients developed MAS similarly to the *hemophagocytic lymphohistiocytosis* (HLH) syndrome [13,18–20]. The HLH is characterized by an exacerbation of innate responses with increases in the CRP levels, ferritin, fibrinogen, D-dimer and interleukin-1 (IL-1) and IL-6 [21]. It is immediately treated via pulse therapy with corticosteroids, anticoagulants, broad-spectrum antibiotics in order to prevent superinfections, and inhibitors of interleukins and immunoglobulins [19,21,22].

Regarding SARS-CoV-2, IL-6 has been used for the characterization of the cytokine storm in severe COVID-19; however, hypoxemia has been the criterion that helps to determine the administration of corticosteroids. Historically, corticosteroids have been effective in immunomodulating hyperinflammation in patients with severe acute respiratory syndrome coronavirus (SARS-CoV) and Middle East respiratory syndrome coronavirus (MERS-CoV). However, the use of corticosteroids, both in COVID-19 and in the SARS and MERS epidemics, has become a great conundrum, since the early use led to adverse consequences such increased viral load and worsening of the disease, but when properly administered, corticosteroids lead to an amelioration, such as reduction of fever, improvement of oxygenation and reduction of mortality rate [2,23,24]. Even so, the appropriate timing and dosage for their administration still represent a dilemma in which there is a choice between uncontrolled viral infection versus reduced progressive deterioration of oxygenation indicators, rapid imaging progress and excessive inflammatory response [23,25].

Thrombotic complications are also an important issue in COVID-19 [26–33]. Clinical and laboratory findings of COVID-19 patients included thrombocytopenia, elevated D-dimer, prolonged prothrombin time, and disseminated intravascular coagulation [33,34]. In addition, elevated D-dimer levels and hyperfibrinogenemia were used as criteria for the application of an aggressive thromboprophylaxis [31]. Still, co-infections involving respiratory bacteria and viruses are commonly identified as important causes of morbidity and mortality [35,36]. In COVID-19, secondary bacterial infections are more common than co-infections, especially in patients developing severe COVID-19 [37]. CRP is often used as a solution for a clinical dilemma. Although it is an acute phase protein in response to inflammation, it is the only point-of-care biomarker of bacterial infection used to determine antibiotic treatment in patients with acute respiratory infections as well as for monitoring antibiotic treatment in patients with known bacterial infections and for differentiating between bacterial and viral infections in various clinical situations [38–41].

In the present study, we will demonstrate how the early and longitudinal monitoring clinical-based practice, as well as quantifying 5 markers (CRP, IL-6, fibrinogen, ferritin, D-dimer) daily has served to reduce the rates of mortality due to COVID-19 among the health professionals from Porto Velho, the capital of the Rondônia state. This longitudinal follow-up of inflammatory markers subclassified patients among those who needed only antibiotic therapy (defined here as YELLOW PATIENT); those in which antibiotic therapy and anticoagulant therapy were sufficient for resolution of COVID-19 (RED PATIENT); and the third group in which COVID-19 evolved into a severe disease (PURPLE PATIENT), and for which antibiotic therapy, anticoagulant therapy and pulse therapy administration resolved the infection and prevented the progressive deterioration of lung injury. To this end the observation of CRP and IL-6 kinetics levels can be markers to distinguish the timing of the cytokine storm in patients with a more severe form of COVID-19 in relation that one that presented COVID-19 with endothelial injury determining the therapy to be conduct.

## Materials and Methods

### COVID-19 severity early identification plan

The state of Rondônia instituted the Home Medical Care Service (HMCS) more than 10 years ago to treat patients with chronic diseases via home care, to relieve overcrowding the public hospitals in the state network. With the emergence of the SARS-CoV-2 pandemic, this service was intensified in view of the high mortality rates of health professionals due to fight against COVID-19. The principal goal was the early identification of cases followed by a severity analysis and follow-up based on daily monitoring of clinical-laboratory parameters to assess the patient’s vulnerability potential. This follow-up was carried out by the HMCS and consisted of a nursing team responsible for blood sample collection, monitoring of vital signs and medicine administration when necessary, and a medical remote health team that, when necessary, included a physical visit. The Rondonia State Laboratory of Pathology and Clinical Analysis (LEPAC) supplied the kits for measure the markers used for the follow-up of the patients. The State Coordinating Office for Management and Pharmaceutical Assistance carried out the provision of supplies and medicines to avoid shortages and enable an uninterrupted flow of administration to the patients. Laboratory tests were performed daily by LEPAC.

### Clinical and laboratory monitoring of patients with COVID19

Health professionals as well as military personnel and their respective family members were referred to the Oswaldo Cruz Polyclinic (OCP) where they were submitted to a medical evaluation and diagnoses of SARS-CoV-2 infection by RT-PCR, via intranasal swab sample collection, or rapid serological test. There was a total of 8,902 visits to the Oswaldo Cruz Polyclinic (OCP) in the period from 2^nd^ June 2020 to 31^st^ December 2020. Of this, 631 patients were followed up by HMCS due to clinical laboratory markers alterations. Within this group of patients, we had four deaths. Mortality of patients cared for by the HMCS was 0.045%.

The methodology used for the running of HMCS follows: the medical evaluation screened the patients for vital signs and the severity of infection. All patients who had no advanced symptoms or signs of clinical deterioration were referred for laboratory markers quantifying at LEPAC. Depending on the results of this laboratory markers and the clinical condition, daily follow-up was conducted involved laboratory markers new determinations’ when necessary. The treatment was also personalized at home by a HMCS nursing team followed by checking vital signs and administering medication. The medical team carried out the remote monitoring and, when necessary, carried out a physical visit.

The study was approved by the Fundação de Medicina Tropical Dr. Heitor Vieira Dourado Ethics Review Board (CAAE: 51963021.9.0000.000) and the requirement for informed consent was waived by the Ethics Commission because these data come from medical and laboratory assistance that took place between 2020 and 2021 and medical records were available anonymously encoded.

### Screening of patients

During the medical consultation, patients who presented signs of clinical severity evidenced by a Glasgow score of ≤14, respiratory rate above 24 respiratory incursions per minute (RIPM) and O2 saturation less than 90% were referred for hospitalization to receive therapeutic and ventilatory support or admission to the ICU. If the patient had a respiratory rate between 21 and 24 RIPM or alterations in any of the laboratory markers such as CRP, fibrinogen, D-dimer, ferritin and IL-6, the patient was immediately followed up at home on a personalized and daily basis by the HMCS.

### Study population and risk classification

All patients were grouped according to the clinical signs and symptoms and laboratory markers in BLUE, GREEN, YELLOW, RED or PURPLE patient as follows:

#### BLUE PATIENT

Patients that do not present alterations in laboratory markers and were classified as having a viral syndrome. All these patients were instructed to take 10,000 U vitamin D every 48 h due to hypovitaminosis of vitamin D being described in patients with severe COVID-19 [42]. In addition, 600 mg of acetylcysteine every 12 h were prescribed due to its glutathione-forming potential, which protects against oxidation by iNOS [43].

#### GREEN PATIENT

characterized as patients that only presented increased ferritin levels. For these patients, a therapeutic based on iron metabolism to decrease the uptake and absorption of iron [44] was prescribed, as well as vitamin D and acetylcysteine as mentioned above. The therapy was composed of i) 60 mg of chelated zinc as an inhibitor that competes with serum iron ion; ii) proton pump inhibitor 2 h before lunch to control gastric acidity to prevent the transformation of ferric ions into ferrous ions absorbed in the intestine; iii) restriction of red meat intake because it is the main source of iron.

#### YELLOW PATIENT

Patients showed elevated CRP in the first examinations, without increases in fibrinogen. These patients required only antibiotic therapy and CRP levels served as a guide [41]. If an increase in CRP greater than 5 and less than 20 mg/L, more than 24 RIPM and/or fever, antibiotic therapy with amoxicillin clavulanate for 7 days was provided. CRP greater than 20 mg/L were given venous antibiotic therapy with amoxicillin clavulanate with levofloxacin or oral doxycycline.

#### RED PATIENT

Include the ones who presented significant increases in CRP and IL-6 concomitantly, and principally due to alterations in fibrinogen. These patients received antibiotic therapy with amoxicillin clavulanate and 1 mg/kg/day of enoxaparin. They were characterized as having a severe endothelial lesion due to increased fibrinogen even after 1 mg/kg/day of enoxaparin for more than 48 h.

#### PURPLE PATIENT

Initially all were classified as RED PATIENTS but evolved to cytokine storm syndrome. Some of them responded with alterations in CRP, ferritin, and IL-6, in addition to fibrinogen. Even when after returning to normal levels, these patients presented proportionally higher IL-6 levels than CRP levels, which indicated an exacerbation of cytokines, and fibrinogen rose again after a few days. For these patients, in addition to antibiotic coverage, the treatment conducted with 2 mg/kg/day of enoxaparin, deworming with ivermectin to avoid the risk of strongyloidiasis infestation, and pulse therapy with 20 mg dexamethasone on the 1^st^ day followed by 3 days of 10 mg/day and 3 days of 5 mg/day.

## Results

### Subclassification of patients based on follow-up of the five laboratory markers

Six hundred thirty-one patients had clinical laboratory alterations and were grouped according to the clinical and laboratory symptoms described above. To demonstrate biochemical and hematological parameters in the first examination a sampling of the total patients was used (Table 1) such as twenty-seven of BLUE PATIENT group; twenty-eight of YELLOW PATIENT group; twenty-three of RED PATIENT group and thirty-two of PURPLE PATIENT group. In addition, a group of 27 cases, between 30 and 55 years-old, with a RT-PCR negative diagnosis of COVID-19 and no symptoms was recruited to be followed up as a CONTROL group. The analysis did not show differences between patients belong to either YELLOW, RED or PURPLE groups, little changes in the neutrophil/lymphocyte ratio and in TGO and TGP levels were observed in PURPLE group. To this end, this first examination follow-up cases with the five laboratory markers was important for the subclassified patients.

**Table 1.** Biochemical and hematological parameters in the first examination

|  | CONTROL<br>N=27 | BLUE PATIENT<br>N=33 | YELLOW PATIENT<br>N=28 | RED PATIENT<br>N=23 | PURPLE PATIENT<br>N=32 | p |
| --- | --- | --- | --- | --- | --- | --- |
| gender (% male). $\chi^2$ | 44.4 | 48.5 | 32.1 | 43.5 | 71.9 * | 0.0343 |
| age | 35 (24.2 ; 42.2) <sup>a</sup> | 33 (26.5 ; 44.5) <sup>a</sup> | 40 (33.0 ; 48.5) <sup>ab</sup> | 44.5 (35.2 ; 54) <sup>ab</sup> | 46 (39.7 ; 62) <sup>b</sup> | 0.0002 |
| Hematocrit | 38 (36.18 ; 40.15) | 39.35 (38.75 ; 41.58) | 38.8 (37.05 ; 41.9) | 37.8 (35.9 ; 40.35) | 43.6 (37.35 ; 46) | 0.0562 |
| Hemoglobin | 13.5 (12.28 ; 14.2) | 13.8 (12.93 ; 14.8) | 13.75 (12.7 ; 15) | 13.2 (12.05 ; 13.88) | 15.1 (12.6 ; 16.4) | 0.0952 |
| Red Blood cells | 4.4 (4.245 ; 4.988) | 4.71 (4.46 ; 5.04) | 4.925 (4.485 ; 115.7) | 4.66 (4.433 ; 304.3) | 5.1 (4.55 ; 5.325) | 0.0936 |
| VCM | 84.75 (83.35 ; 87.65) | 85.5 (82.73 ; 87.48) | 85.2 (82.35 ; 87.53) | 85.1 (82.88 ; 88.23) | 87.3 (83.8 ; 88.5) | 0.7592 |
| HCM | 29.95 (28.65 ; 30.9) | 29.45 (28.28 ; 30.83) | 29.65 (28.95 ; 30.35) | 29.45 (27.68 ; 30.3) | 30.4 (28 ; 31.7) | 0.7404 |
| CHCM | 35.05 (34.3 ; 35.53) | 34.9 (33.35 ; 35.53) | 34.45 (33.95 ; 34.93) | 34.35 (33.7 ; 35.28) | 34.3 (32.95 ; 35.65) | 0.4266 |
| Leukocytes | 7400 (6675 ; 9060) | 6635 (5253 ; 9833) | 6035 (3340 ; 7853) | 6485 (5060 ; 9638) | 7870 (5850 ; 9670) | 0.1909 |
| Eosinophils | 147.4 (81.7 ; 381.2) | 133.9 (63.95 ; 212.9) | 83.3 (0 ; 242.2) | 57.7 (0 ; 136.1) | 138.0 (80 ; 270.6) | 0.0670 |
| Neutrophils | 4056 (2698 ; 4829) | 3396 (1778 ; 5002) | 2816 (1490 ; 4486) | 3893 (3083 ; 6118) | 5180 (3293 ; 6705) | 0.1064 |
| Lymphocytes | 2172 (1492 ; 2755) | 2315 (1216 ; 2906) | 2045 (1650 ; 2689) | 1928 (1518 ; 2640) | 1754 (1224 ; 2640) | 0.8516 |
| Monocytes | 427 (311.6 ; 621.3) | 426.7 (197.7 ; 627.1) | 407.4 (271 ; 642.2) | 554.4 (320 ; 624.8) | 483 (205.7 ; 646.4) | 0.9063 |
| Platelets | 243500 (173750 ; 333250) | 252000 (209500 ; 315250) | 226500 (186250 ; 273250) | 252500 (181750 ; 290000) | 219000 (166500 ; 294000) | 0.4995 |
| Neu/Lymp ratio | 1.902 (1.404 ; 3.007) <sup>ab</sup> | 1.53 (1.225 ; 3.074) <sup>ab</sup> | 1.27 (0.8698 ; 1.922) <sup>a</sup> | 1.813 (1.444 ; 3.295) <sup>ab</sup> | 2.957 (1.339 ; 3.652) <sup>b</sup> | 0.0153 |
| Lymp/Mono ratio | 4.5 (3.216 ; 5.544) | 5 (3.833 ; 6.25) | 5.3 (3.771 ; 6.167) | 4.4 (2.888 ; 5.065) | 4.25 (2.795 ; 6.875) | 0.4992 |
| PLT/Lymp ratio | 109.7 (79.41 ; 174.7) | 124.2 (82.05 ; 182.7) | 103.1 (93.33 ; 130.8) | 125.3 (100.7 ; 153.9) | 118.4 (76.33 ; 155.1) | 0.8079 |
| urea | 22.4 (20.05 ; 30.25) | 23.6 (20.33 ; 31.68) | 22.2 (18.5 ; 26.6) | 25.3 (18.1 ; 32.28) | 36.4 (21.5 ; 38.9) | 0.1946 |
| creatinine | 0.9 (0.6225 ; 1.045) | 0.9 (0.745 ; 1.135) | 0.8 (0.765 ; 1.103) | 0.7 (0.6125 ; 0.925) | 0.9 (0.82 ; 1.075) | 0.1223 |
| sodium | 140.5 (138 ; 141.5) | 137.0 (134.8 ; 141.3) | 140.0 (139 ; 141) | 139.5 (138 ; 141.5) | 138.5 (135.5 ; 141.5) | 0.2044 |
| potassium | 3.9 (3.668 ; 4.173) | 3.8 (3.798 ; 4.423) | 3.9 (3.645 ; 4.255) | 4.0 (3.94 ; 4.38) | 4.0 (3.833 ; 4.453) | 0.7590 |
| TGO | 19.5 (15.75 ; 26.25) | 22.0 (19 ; 30) | 30.0 (19.45 ; 60.5) | 31.0 (19 ; 60.25) | 22.0 (18 ; 29) | 0.0464 |
| TGP | 17.5 (12 ; 28.5) <sup>a</sup> | 26.0 (15 ; 48) <sup>ab</sup> | 32.3 (19 ; 67.25) <sup>ab</sup> | 36.5 (21 ; 74) <sup>b</sup> | 29.0 (22 ; 40.5) <sup>ab</sup> | 0.0445 |

Median (25 ; 75 interquartiles ). Anova Kruskal Wallis and Dunn's multiple comparisons test

The YELLOW PATIENT group showed CRP elevations, and the use of antibiotics decreased continuously these levels until the baseline (Figure 1A). In these patients, IL-6 levels behaved more timidly and decreased synchronously with CRP levels (Figure 1B). Fibrinogen levels did not exceed the limit of 437 mg/dL despite the mild increase in IL-6 levels (Figure 1C) and, therefore, the use of enoxaparin was not necessary. Without increased in fibrinogen levels, there was no thrombus formation and, therefore, the D-dimer, a predictor of fibrinolysis, did not increase (Figure 1D). Figure 1E shows the ferritin curve in women. The variation showed that some female patients demonstrated iron metabolism disorder, requiring treatment according to the initial protocol. Antibiotic treatment led to decreased CRP and IL-6 levels. Figure 1F demonstrated the ferritin levels in men. In most of the male patients, ferritin levels were much higher than in women in the first examination, which is probably due to the eating habits of men. Similarly, the treatment of iron metabolism and control of metabolic syndromes resulted in optimal results, with the decrease of ferritin levels and the reduction of CRP and IL-6 levels concomitantly. These patients had mild to moderate clinical symptoms, anosmia, cough, fever, and chest discomfort, and they did not require the oxygen support.

**Figure 1.**
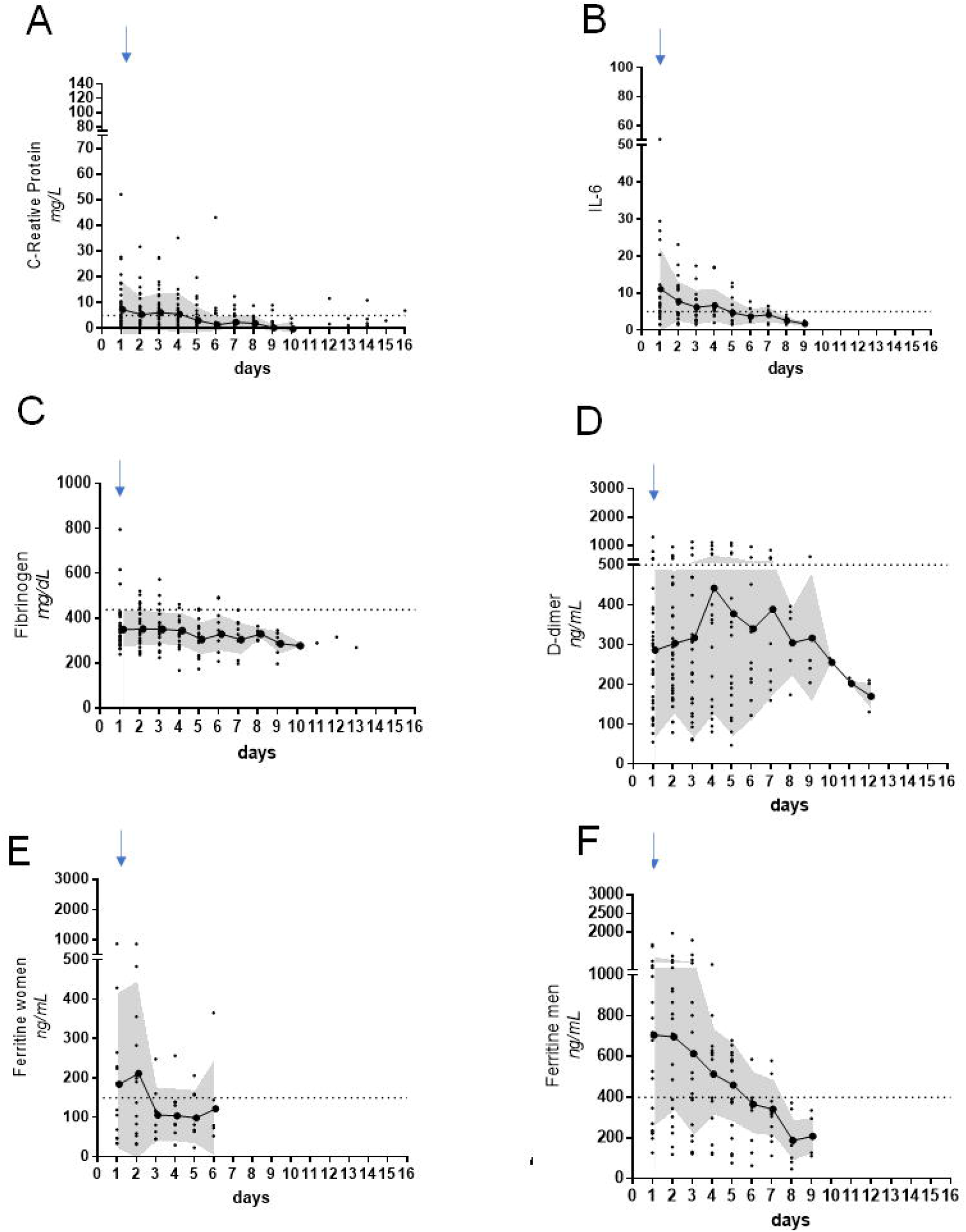
Kinetics of inflammatory markers in YELLOW PATIENTs. The behavior of the markers in patients whose antibiotic therapy with amoxicillin clavulanate was provided for 7 days. A) CRP levels during 16 days of follow up; B) IL-6; C) Fibrinogen; D) The D-dimer; E) The ferritin curve in women; F) The ferritin levels in men. Blue thin arrows indicate the beginning of antibiotic therapy. The line represents the mean of data and gray area the standard deviation.

Figure 2 represents the behavior of the markers in RED PATIENTs who received venous antibiotic therapy and subcutaneous anticoagulant and evolved to improvements in clinical signs and laboratory results. These patients were treated with 1 mg/kg/day of enoxaparin which resulted in reversal of fibrinogen levels to normal titers. CRP started with higher levels and, when patients were treated with venous antibiotic therapy, they had progressive resolution (Figure 2A). The increase in IL-6 levels accompanied the increases in CRP, but when CRP levels were below 25 mg/L, with the venous antibiotic therapy there was a gradual decrease in IL-6 to normal levels (Figure 2B). In Figure 2C it is possible to observe how increased fibrinogen was used within this protocol as the standard for anticoagulant use (see red arrow). The graph demonstrated that after the increase of fibrinogen levels above 437 mg/dL, the indication of a daily dose of 1 mg/kg/day of enoxaparin for nine days was sufficient for the normalization of the results. In Figure 2D it is presented the increase in D-dimer levels on the second day of examination demonstrating that these patients had already had previous increases in fibrinogen, since the peak that occurred on the fifth day of examinations represented the increase in fibrinogen on the second day (Figure 2C). After this, the D-dimer curve showed that the patient no longer had fibrinolytic activity after anticoagulant therapy. In this group, female patients already showed higher levels of ferritin (Figure 2E). Regarding the male patients, most of them also had alterations associated with metabolic syndromes (Figure 2F).

**Figure 2.**
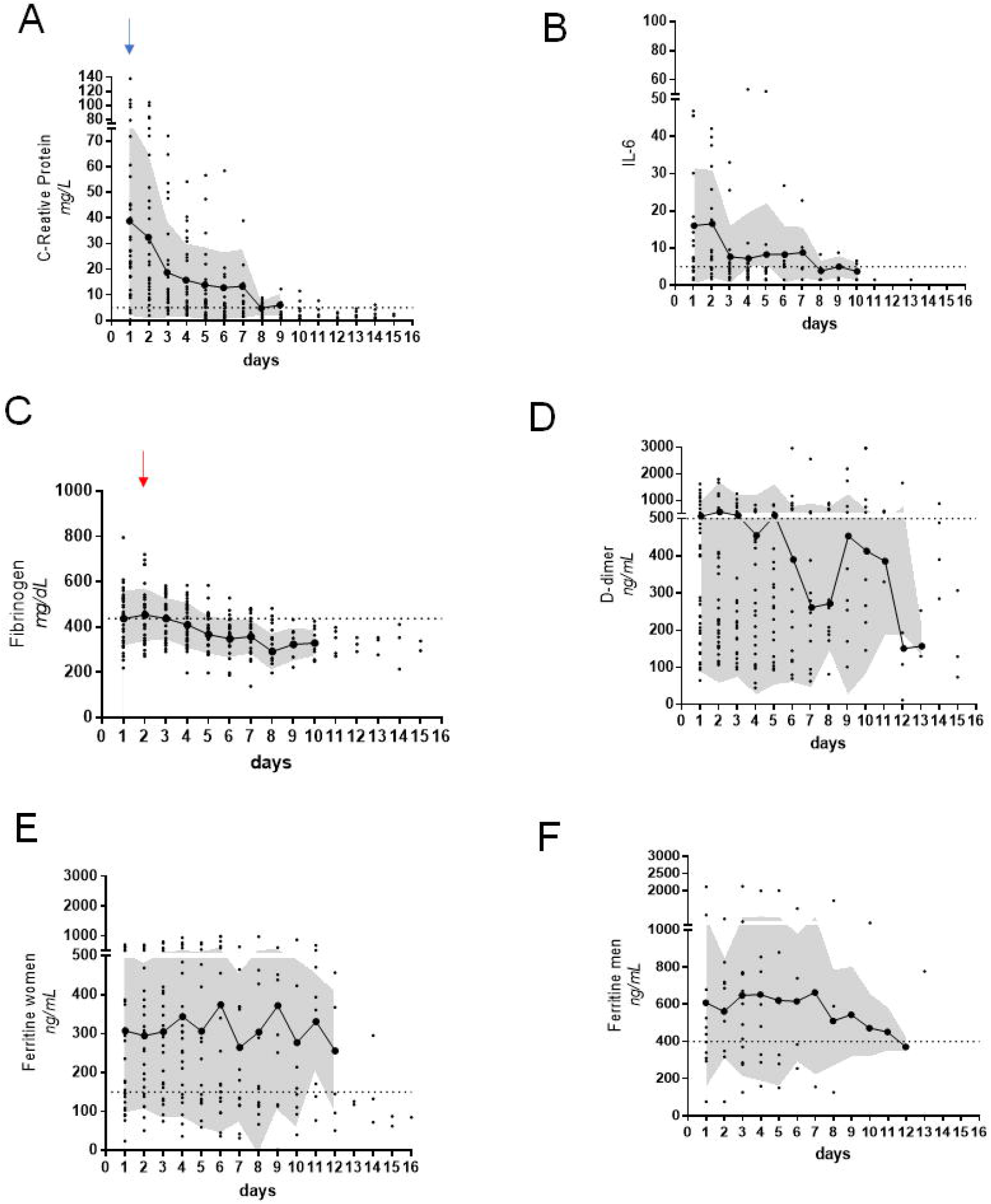
Kinetics of inflammatory markers in RED PATIENTs. The behavior of the markers in patients treated with venous antibiotic and anticoagulant. A) CRP levels during 16 days of follow up; B) IL-6; C) Fibrinogen; D) The D-dimer; E) The ferritin curve in women; F) The ferritin levels in men. Blue thin arrows indicate the beginning of antibiotic therapy. Red thin arrows indicate the beginning of anticoagulant therapy. The line represents the mean of data and gray area the standard deviation.

Figure 3 describes the behavior of the markers of patients who underwent an abrupt increase in IL-6 levels, which is the main marker for the characterization of a cytokine storm. In Figure 3A, CRP levels in these patients begun with high levels and, after the start of antibiotic therapy, there is a progressive drop. Between sixth-eighth day of illness, in patients with the most severe symptoms, IL-6 levels increased earlier than those of CRP levels, and the peak of IL-6 in pg/mL is proportionally higher than the peak of CRP in mg/L (see thick green arrow, Figure 3B). This abrupt increase characterizes the cytokine storm, which was treated with high-dose corticosteroids and resulted in a significant drop in the release of IL-6 until they reach normal levels. This drop is also observed with CRP levels. In Figure 3C, in the graph showing the fibrinogen levels it is clearly demonstrated that when the patient was recruited by the service, the patient already presented increased levels of CRP, IL-6 and fibrinogen levels (see thin red arrow), which demonstrates that these patients had already had the disease for several days. The introduction of antibiotic therapy with enoxaparin at 1 mg/kg correlates with the drop of fibrinogen to normal levels, but the sharp increase in IL-6 levels, which denotes a cytokine storm, creates a new increase in fibrinogen levels that is characterized as a second wave, even when using 1 mg/kg of enoxaparin. The re-appearance of an elevation in fibrinogen was an indicator for an increase in anticoagulation therapy with enoxaparin of 2 mg/kg day (see blue and red thick arrows, Figures 3A and C). This results in the start of the decrease of these markers to normal levels. In Figure 3D, the first increase in D-dimer levels occurred a few days after the peak of fibrinogen levels, around the eighth to tenth day. The new increase in D-dimer occurred on the thirteenth day, which was after the second peak of fibrinogen levels that was observed between the eighth to the tenth day (Figure 3C). In Figure 3E, the increase in ferritin levels in female patients shows that these patients have had the disease for some days, and from the sixth day, there is an exponential increase and the patient’s improvement response is translated into the decrease in the curve. Figure 3F showed the increase in ferritin levels in male patients, with an expected upward and progressive curve as can be seen in infections, but around the sixth to seventh day there is a greater than 80% increase in relation to the previous day, demonstrating that ferritin is an excellent marker of inflammatory response. Only patients who developed this abrupt increase compared to the previous day were treated with corticosteroid via pulse therapy. The observation of CRP and IL-6 kinetics levels allowed us to distinguish the timing of the cytokine storm in patients with a more severe form of COVID-19 in relation that one that presented COVID-19 with endothelial injury. Although the units of CRP and IL-6 are different, at the beginning of the disease the kinetics of CRP and IL-6 levels in these two groups of patients are similar, though with the peak IL-6 in pg/mL being higher than the peak of CRP in mg/L (Figures 4A and B).

**Figure 3.**
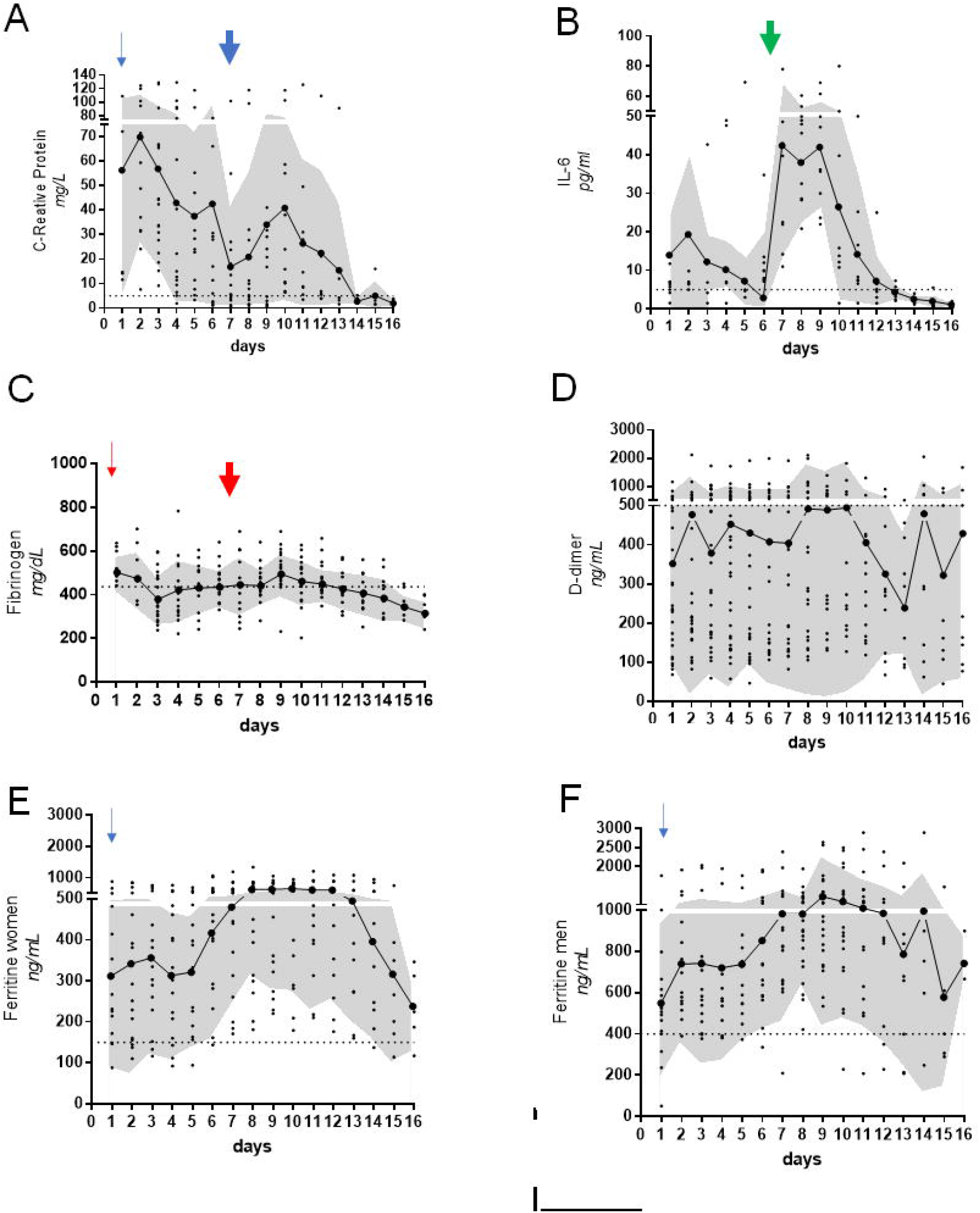
Kinetics of inflammatory markers in PURPLE PATIENTs. The behavior of the markers in patients treated with venous antibiotic and anticoagulant. A) CRP levels during 16 days of follow up; B) IL-6; C) Fibrinogen; D) The D-dimer; E) The ferritin curve in women; F) The ferritin levels in men. Blue thin arrows indicate the beginning of antibiotic therapy. Red thin arrows indicate the beginning of anticoagulant therapy. Blue thick arrow indicates broad-spectrum antibiotic therapy in order to prevent superinfections; Red thick arrow indicates the increase in anticoagulation therapy with enoxaparin of 2 mg/kg day; Green thick arrow indicates the pulse therapy due the inversion of IL-6 levels in relation to CRP for the prediction of the cytokine storm. The line represents the mean of data and gray area the standard deviation.

**Figure 4.**
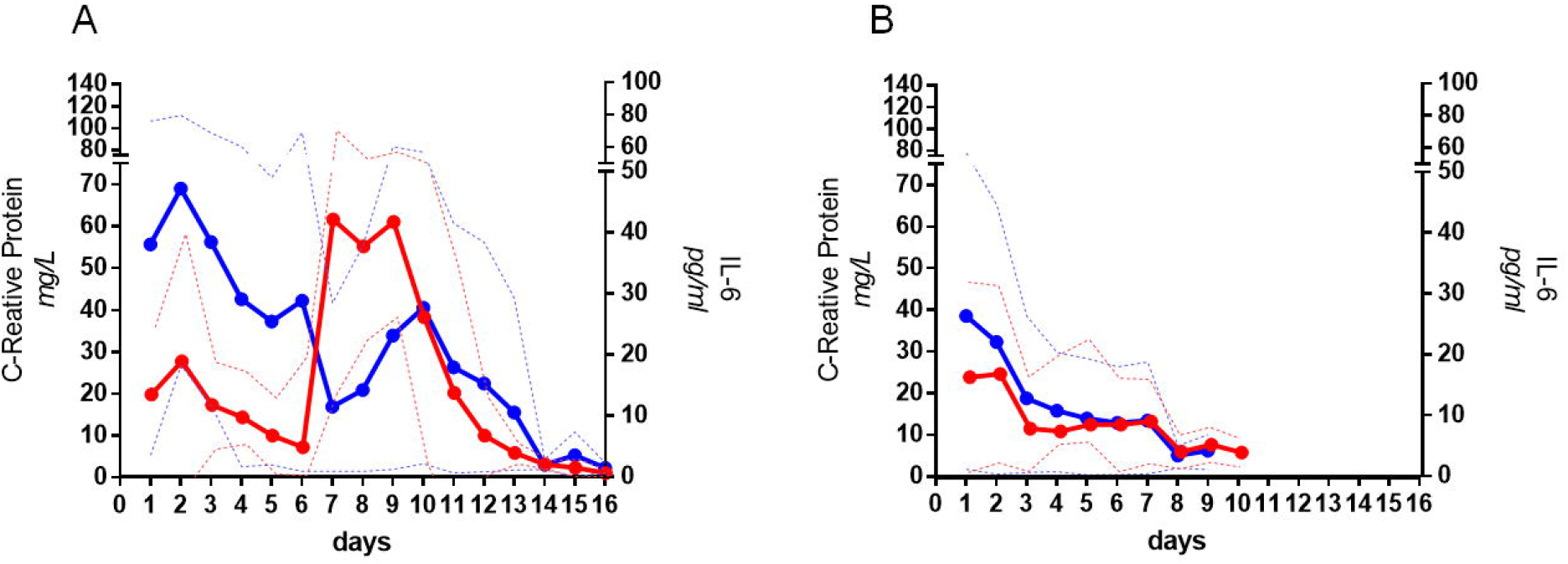
Comparison of the kinetics of CRP and IL-6 in PURPLE and RED PATIENT groups. CRP and IL-6 kinetics allow distinguish the timing of the cytokine storm in PURPLE PATIENTS. Although the units of CRP and IL-6 are different, at the beginning of the disease in these two groups of patients are similar, though with the peak of CRP in mg/L being higher than the peak IL-6 in pg/mL. The thick lines represent the mean of data and dashed lines the standard deviation, RED the IL-6 kinetics and BLUE CRP kinetics. A) PURPLE PATIENTs; B) RED PATIENTs.

### Dynamics of laboratory markers in the determination of therapy

The dynamics of serological markers allowed us to determine the antibiotic therapy, the anticoagulant therapy, and the administration of pulse therapy. Patients defined as having bacterial co-infection had their infection resolved only with antibiotics. IL-6 and CRP levels served as a guide for the choice of the spectrum of the antibiotics. These patients showed CRP alterations of between 5.0 and 20.0 mg/L, and IL-6 up to 30 pg/mL and received antibiotic therapy (ABT) of 875 mg amoxicillin and 125 mg potassium clavulanate every 12 h for 7 days (Figure 5A). Fibrinogen also served as a guide for treatment; if the fibrinogen levels were less than 437 mg/dL, they continued to receive antibiotic therapy. Those patients who had ferritin alterations were also prescribed vitamin D, acetylcysteine and received anti-iron uptake therapy. The other markers were unaltered, and the infection was resolved in these patients.

**Figure 5.**
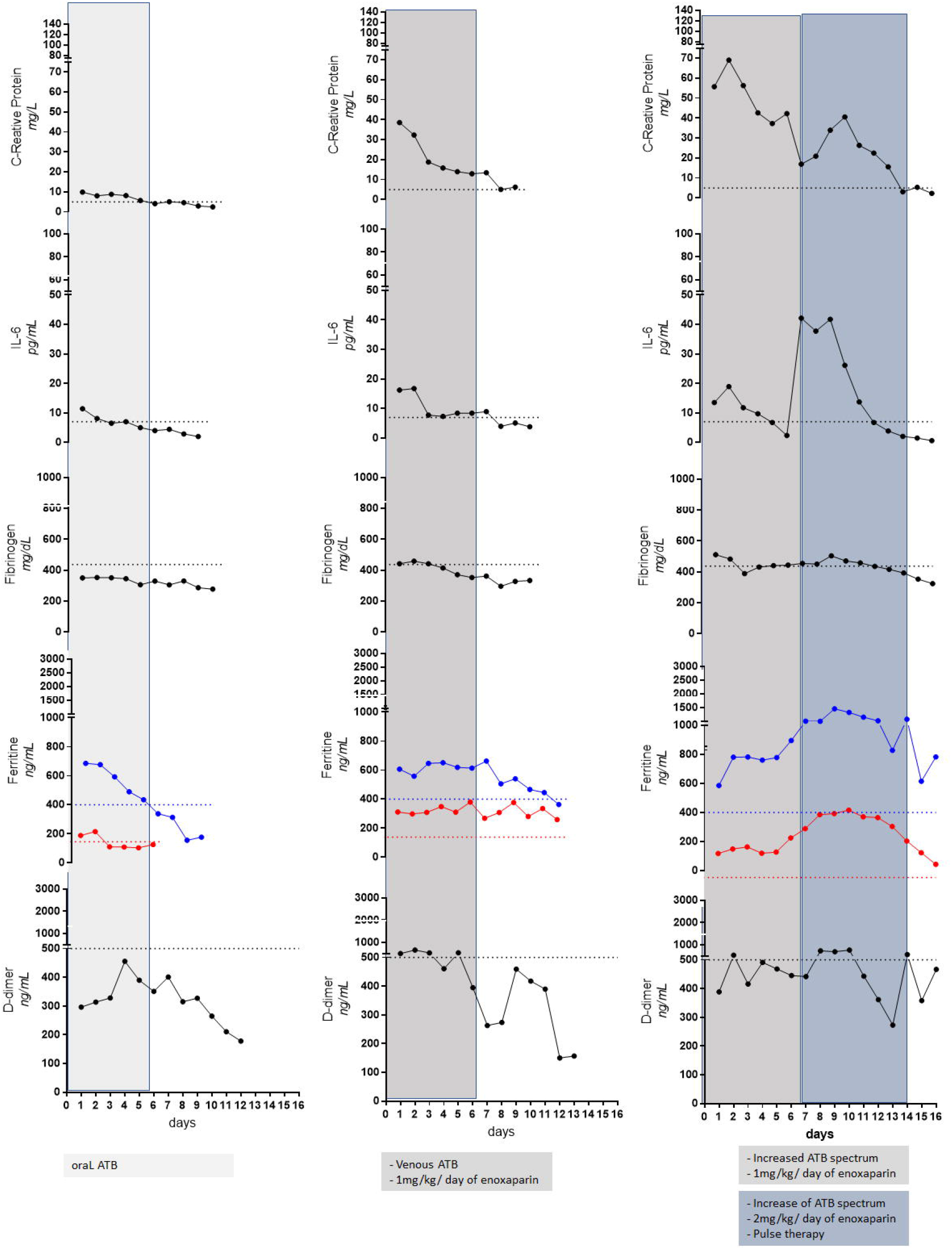
The determination of therapy based on the dynamic of laboratory markers. A) YELLOW PATIENTs had their infection resolved only with antibiotics. These patients showed CRP alterations of between 5.0 and 20.0 and IL-6 up to 30 pg/mL and received antibiotic therapy (ABT) of 875 mg amoxicillin and 125 mg potassium clavulanate every 12 h for 7 days. The other markers were unaltered, and the infection was resolved in these patients. B) RED PATIENTS showed increases in CRP levels to over 20 and IL-6 up to 30 pg/mL and concomitant increases in fibrinogen to over 437. For the treatment of opportunistic, ABT was replaced by 1 g of cefepime via venous route and 500mg clarithromycin via oral route every 12 h, and patients received enoxaparin at 1 mg/kg/day subcutaneously for 7 days. C) PURPLE PATIENTs showed CRP levels to over 20 and IL-6 up to 30 pg/mL and concomitant increases in fibrinogen to over 437. For the treatment of opportunistic infections, ABT was replaced by 1 g of cefepime via venous route and 500mg clarithromycin via oral route every 12 h, and patients received enoxaparin at 1 mg/kg/day subcutaneously for 7 days. CRP and IL-6 kinetics distinguished the timing of the cytokine storm with the peak IL-6 in pg/mL being higher than with the peak of CRP in mg/L. The pulse therapy was given with dexamethasone 20 mg on Day 1, followed by 3 days at 10 mg/day and 3 days of 5 mg/day. They received an increase in the range of coverage of antibiotics, with the addition of meropenem and amikacin, or tazobactam. The dose of enoxaparin was increased to 2 mg/kg/day since fibrinogen rose again after a few days even after 1 mg/kg/day of enoxaparin for more than 48 hours. The line represents the mean of data.

Patients with endothelial injury showed increases in CRP levels to over 20 mg/L and IL-6 up to 30 pg/mL and a concomitant increase in fibrinogen to over 437 mg/dL (Figure 5B). For the opportunistic infection’s treatment, ABT was replaced by 1 g of cefepime via venous route and 500 mg clarithromycin via oral route every 12 h, and patients received 1 mg/kg/day of enoxaparin subcutaneous for 7 days. If ferritin levels were high, in addition to vitamin D, acetylcysteine and anti-iron uptake therapy was administered. The patients we also investigated for type 2 diabetes mellitus, hepatic steatosis, and drug hepatitis, in addition to renal function. The infection was resolved with antibiotics and CRP levels were decreased in these patients. Treatment with 1 mg/kg/day of enoxaparin decreased fibrinogen in 24 h and therefore there was no increase in D-dimer. Initially, it was not observed patients with increased fibrinogen without increased CRP levels, and this endothelial lesion usually came from a mixed infection. Increased fibrinogen production behaved as a marker of the onset of endothelial injury which results from the exposure of tissue factor in the basal membrane (extrinsic via the coagulation cascade with subsequent platelet adhesion).

Patients with cytokine storm syndrome showed increased fibrinogen even after 1 mg/kg/day of enoxaparin for more than 48 h and, even when after returning to normal levels, fibrinogen levels increase again after a few days (Figure 5C). Unlike patients with endothelial injury, increased fibrinogen occurred concomitantly with CRP, IL-6 and ferritin levels. The follow-up of ferritin was very important, since the sharp increase in its values between 70 and 100% of the initial levels together with IL-6 peaks caused an exacerbation of cytokines, indicating macrophage activation syndrome. In addition, patients received therapy to correct metabolic syndrome and iron metabolism. These patients were characterized as presenting severe endothelial lesion and the parameters observed guided the treatment of a possible macrophage activation syndrome or even hematophagous lymphohistiocytic syndrome. After deworming with nitazoxanide to avoid the risk of strongyloidiasis infestation, these patients received an increase in the range of coverage of antibiotics, with the addition of meropenem and amikacin, or tazobactam, and the dose of enoxaparin was increased to 2 mg/kg/day, and pulse therapy was conducted with 20 mg of dexamethasone on Day 1, followed by 3 days of 10 mg/day and 3 days of 5 mg/day (Figure 5C).

## Discussion

There are no data on the literature demonstrating a home care follow-up of markers in COVID-19 patients [23,45,46]. Our study is the first to show a clinical and laboratory follow-up of patients with COVID-19 since the first symptoms conduct in home care. In a sampling of the presentation of COVID-19 disease, we observed this heterogeneity from asymptomatic cases to patients who were evolving to SARS, as observed everywhere [3,47–49]. Some patients required oxygen therapy, but daily follow-up prevented worsening and the mortality rate was very low. Patients who needed oxygen fell into RED and PURPLE groups. Even with the increase of the markers levels after treatment, only some RED PATIENTS still needed oxygen support. In PURPLE PATIENTs, the computed tomography scans have already showed a high rate of lung involvement (personnel communications of clinicians from HMCS) and all patients needed oxygen support. Moreover, the subclassification and monitoring of the markers evaluated allowed us to understand the dynamics of the pathophysiology of SARS in severe COVID-19, which is usually characterized as a cytokine storm [1–5]. We had patients who presented the cytokine storm syndrome and, with very rare exceptions; surprisingly none of the patients evolved for the necessity of mechanical ventilation or were at risk of death, as commonly observed [2,3,6–10].

The follow-up was completely effective regardless of a first consultation, or even without a tomographic image as an effective marker of severity. These observations led us to create a therapeutic protocol associated with the clinical picture: CRP as a guide for the introduction of antibiotics, fibrinogen as a guide for the introduction of anticoagulation therapy with enoxaparin and, IL-6 as a guide for the introduction of pulse therapy, which were effective for identifying the cytokine storm [29]. Thus, our greatest finding was that the administration of broad-spectrum antibiotic therapy and pulse therapy with dexamethasone and an increased dose of anticoagulant which contributed to resolve the infection of the group of patients who had a COVID-19 of greater severity and were at risk of death.

Moreover, our study had the opportunity to follow-up all the LEPAC results from the onset of symptoms and the alterations of the evaluated markers gave us clues to the worsening of the disease. The follow-up led us to believe that the markers have a sequence of increases such as: CRP was the first to increase in patients without symptoms, and it increased with the onset of symptoms accompanied by IL-6 and ferritin levels. With the use of antibiotic therapy, CRP dropped substantially in the YELLOW PATIENTs with a good therapeutic response, improvement in the clinical presentation and laboratory markers in 48 h.

CRP could be a marker of opsonization at the first sign of a primary bacterial infection, as well as the secondary one resulting from an infectious respiratory viral disease, such as COVID-19 [37,41]. This fact makes us rethink the relationship of CRP with the virus, because CRP would not decrease if it was only associated with inflammation, whereas the association with high CRP being most commonly associated with a secondary bacterial infection by respiratory viruses [38–40,50,51]. Although prescribing broad-spectrum antibiotics is important for preventing these infections, the use of antibiotics should be prudent during the evolution of the SARS-CoV-2 pandemic [40,50,52].

In RED PATIENTS, CRP and IL-6 levels were already higher even in the first days of infection, though the patients still present mild symptoms; however, fibrinogen increased soon after. After antibiotic and anticoagulant therapy, the markers improved with resolution of disease. The increase in fibrinogen due to an even greater increase in CRP and IL-6 levels led to the use of antibiotic therapy and anticoagulant treatment that resolved the infection. Our findings call into question the use of fibrinogen as an early marker of this thrombotic stage, as seen in the H1N1 pandemic [35,36]. According to these studies, colonization of the nasopharynx with pathogenic bacteria in respiratory virus infection may predispose patients to a secondary bacterial infection due to impaired mucociliary clearance, thus allowing secondary bacterial invasion, formation of vasculitis zones, capillary thrombosis and necrosis around the areas of bronchiolar damage [35,36].

There are no studies on the pathophysiology of endothelial injury for tissue factor exposure in COVID-19. The D-dimer has been considered one of the severity criteria of SARS, suggesting that disseminated intravascular coagulation leads to death in patients with COVID-19 [31,53]. However, D-dimer levels increased late in relation to fibrinogen, therefore, the alterations in fibrinogen observed here were indicative of the dose of anticoagulant needed in the treatment of COVID-19. Thus, alterations in fibrinogen observed here were resounding in exposing endothelial dysfunction and were indicative of the dose of anticoagulant needed in the treatment of COVID-19.

The PURPLE PATIENTs had behavior similar to RED PATIENT but with high levels markers in the beginning. After antibiotic and anticoagulant therapy, all the markers improved and started to decrease. However, between the ninth and tenth day, some patients had an 80% increase in IL-6 levels from the previous day that was accompanied by a sharp increase of CRP. Although the magnitude of IL-6 pg/mL is different in relation to the CRP mg/L, the peak of IL-6 was earlier and was superior to the peak of CRP (Figure 4). Moreover, these patients had a second wave in fibrinogen evaluation that demanded us to double the dose of the anticoagulant and increase the length of antibiotic therapy in the course of pulse therapy. Thus, our findings bring a new understanding of the sequence of the initial inflammatory response, and no longer place IL-6 as the main protagonist, but the inversion of IL-6 levels in relation to CRP for the role to predict the cytokine storm [54–56].

The cytokine storm plays an important role in the process of aggravation of the disease and is considered one of the main causes of SARS [1,23,56,57]. IL-6 has been responsible for the characterization of the cytokine storm and MAS in severe COVID-19 [11,23,45,54,58]. Effective suppression of the cytokine storm is an important way to prevent the deterioration of patients with severe COVID-19 [59], as has occurred with other epidemics involving SARS, MERS, and severe influenza. The COVID-19 therapy (RECOVERY) randomized controlled and open-label clinical trial showed that use of dexamethasone for up to 10 days resulted in lower mortality over 28 days [60]. The dose used was 6 mg once a day for up to 10 days or until hospital discharge, which is the standard and usual care. The effectiveness of the therapy was low and its benefit was apparently only a mere statistical effect [60], while the use of colchicine in adults hospitalized with COVID-19 was worse [61], due likely severe COVID-19 patients were not separated between who developed endothelial injury like RED PATIENTS from those that developed cytokine storm characterized as PURPLE PATIENTs by the peak of IL-6 followed by CRP. In these patients, their laboratory markers began to demonstrate worsening as a result of the MAS [23,58,62], or even a HLH syndrome [13,18,21,63], prior to the clinical picture, which sometimes did not manifest itself. Our findings are in agreement with Zuñiga and Moreno-Moral teams about high-dose corticosteroid pulse therapy in the increasing of the survival rate of the COVID-19 patients at risk of hyper-inflammatory response [64]. In addition, the pulse therapy would not be sufficient without broad-spectrum antibiotic therapy and the increased dose of cutaneous anticoagulant as reported in several studies [19,21–23,55,58,63].

Ferritin showed a peculiar behavior and was shown to be altered in some patients early in the first exams. Ferritin serves to carry and store iron for vital cellular processes, while protecting against toxicity of this metal for the production of oxygen free radicals [22]. The follow-up of ferritin was very important because a therapy to decrease the uptake and absorption of iron were given to the COVID-19 patients with increased ferritin early on. These patients were clinically well despite having altered ferritin levels due to a pre-existing metabolic syndromes [65,66]. However, this treatment had to be stopped in order not to mask the daily elevation and the increase close to the peak of IL-6 and CRP, as observed in COVID-19 and in syndromes with hyperferritinemia [17,22,55,65,66]. The direct role of ferritin in macrophage activation syndromes, such as COVID-19, lies in the activation of macrophages that play a key role in the perpetuation of hyperferritinemia [17,22,55,65,66]. Herein, patients who had a cytokine storm had an increase of 80 to 100% of ferritin related to the value of the previous day. This fact makes us appreciate the importance of ferritin as an optimal marker of a cytokine storm, especially in those with an initial increase due to metabolic syndrome, and that without prior treatment against absorption/uptake, this would interfere with its reliability.

Our study also has some significant limitations. The drawbacks of the work were that, for some patients there are not all data of five markers daily, despite our efforts to perform the monitoring and dosing of five markers every day. That is due some patients did not present in the LEPAC or they were not found in their residences.

## Conclusions

The home care follow-up was completely effective regardless of a first consultation, or even without a tomographic image, as effective markers of severity. The daily evaluation of the markers was important in order to classify the severity at the onset of symptoms or even when the infection was asymptomatic. CRP as a guide for the introduction of antibiotics, fibrinogen as a guide for the introduction of anticoagulation therapy with enoxaparin and IL-6 as a guide marker for the introduction of pulse therapy. Another observation of the study was the inversion of IL-6 levels in relation to CRP levels to predict the cytokine storm. Moreover, these observations led us to create a therapeutic protocol associated with the clinical picture: This follow-up allowed the patients to be subclassified into: (i) those whose antibiotic therapy was able to resolve the SARS-Cov-2 infection, which was defined as YELLOW PATIENTs; (ii) those whose infection presented endothelial injuries evidenced by increased fibrinogen and which was solved by a higher spectrum venous antibiotic therapy and cutaneous anticoagulant defined here as RED PATIENTs; and (iii) the group of patients with COVID-19 of greater severity in which patients developed a bacterial sepsis and viral response with the cytokine storm and which was resolved with administration of a broad spectrum antibiotic therapy and pulse therapy with dexamethasone and an increased dose of anticoagulant defined as PURPLE PATIENTs.

## Data Availability

All data produced in the present study are available upon reasonable request to the authors

https://data.mendeley.com/datasets/b7kxcyszkh/1

## Author Contributions

SPMMF, FSM and RHM conceived of the study and wrote the initial proposal. SPMMF, FSM, RHM, SAB, LMAC contributed intellectually to the protocol development. PAN estimated the sample size and performed all statistical analysis. SPMMF, FSM, RHM, ABS and DCOM were the lead for the logistics and data collection for the study. FSM, ABS, DCOM, PJG, ELO, JAL, YOC and RSP actively reviewed all data from patients treated by the Multidisciplinary Home Medical Care Service. PJG, ELO, did the laboratory work. The initial draft of the Article was written by PAN and SPMMF, who had full access to and verified all the data underlying the study. All authors had access to the data, agreed to submit for publication, and vouch for the integrity, accuracy, and completeness of the data and for the fidelity of the patients’ laboratory data. SPMMF, JPZ, RHM, LMCA and PAN contributed intellectually to the manuscript. All authors approved the final submitted version.

## Data sharing

Anonymized participant data can be made available upon requests directed to the corresponding author. Proposals will be reviewed on the basis of scientific merit. After approval of a proposal, data can be shared through a secure online platform after signing a data access agreement.

## Acknowledgments

This study had a financial support from Healthy Secretariat of Rondônia State, Brazil. Laboratory supplies, collection supplies and related materials: Fundação Oswaldo Cruz (Fiocruz) - INOVA: VPPCB-005-FIO-20-2-79 and FAPEAM Programa PCTI-EMERGESAUDE Edital N°. 006/2020 (Number 01.02.016301.01078/2021). Projeto financiado com recursos do Programa de Excelência em Pesquisa Básica e Aplicada em Saúde do Instituto Leônidas & Maria Deane – ILMD/Fiocruz Amazônia – PROEP-LABS/ILMD FIOCRUZ AMAZÔNIA We are grateful to all staff, particularly personnel from Multidisciplinary Home Care Service who helped to monitoring and treating of patients. Finally, we thank all of the participants, without whom this research would not have been possible.

